# Modeling social distancing strategies to prevent SARS-CoV2 spread in Israel- A Cost-effectiveness analysis

**DOI:** 10.1101/2020.03.30.20047860

**Authors:** Amir Shlomai, Ari Leshno, Ella H. Sklan, Moshe Leshno

## Abstract

**Objectives:** While highly effective in preventing SARS-CoV-2 spread, national lockdowns come with an enormous economic price. Few countries have adopted an alternative “testing, tracing, and isolation” approach to selectively isolate people at high exposure risk, thereby minimizing the economic impact. To assist policy makers, we performed a cost-effectiveness analysis of these two strategies.

**Methods:** A modified Susceptible, Exposed, Infectious, Recovered and Deceased (SEIRD) model was employed to assess the situation in Israel, a small country with ~9 million people. The incremental cost-effectiveness ratio (ICER) of these strategies, as well as the expected number of infected individuals and deaths were calculated.

**Results:** A nationwide lockdown is expected to save on average 274 (median 124, interquartile range (IQR): 71-221) lives compared to the “testing, tracing, and isolation” approach. However, the ICER will be on average $45,104,156 (median $ 49.6 million, IQR: 22.7-220.1) to prevent one case of death.

**Conclusions:** A national lockdown has a moderate advantage in saving lives with tremendous costs and possible overwhelming economic effects. These findings should assist decision-makers dealing with additional waves of this pandemic.

**Highlights:**

- Drastic measures of national lockdowns are taken by many countries to slow-down SARS-CoV-2 spread. However, these measures have detrimental economic effects.
- Here we compare two strategies to control the epidemic using a modified SEIRD model: 1. Global national lockdown 2. Focused isolation of people at high exposure risk, following detailed epidemiological investigations.
- We show that strategy 1 is modestly superior in saving lives compared to strategy 2, but with tremendous costs to prevent one case of death. This might result in overwhelming economic effects that are expected to increase future death toll.

## Introduction

Since its identification at the very end of 2019, the novel coronavirus (SARS-CoV-2) has spread around the world at an extraordinary rate and has been officially declared a pandemic ^1^. So far, this epidemic has affected over 23 million people and claimed the lives of more than 819,000 individuals at the time of the submission of this manuscript ^2,3^. The first priority in most countries is to enable the local health systems to handle the growing number of patients needing hospitalization and intensive care by “flattening the curve”. Observing the dreadful situation in countries such as Italy and Spain ^4^, many countries are now undertaking extreme measures such as national lockdowns, in effort to stay within the boundaries of the local health system capacity ^5^. However, only a few countries, such as South Korea, have succeeded in slowing the infection rates without enforcing economically damaging lockdowns. Instead, South Korea applied early interventions that included identification and isolation of outbreak sources by massive screening of infected patients and rigorous tracing and isolation of their contacts ^6^. It is not yet clear whether the apparent success of South Korea could be applied to other countries in North America and Europe. It is, however, obvious that extreme non-specific measures are associated with tremendous economic costs and will result in a global financial crisis that will most likely affect public health and other essential aspects of our lives in the coming years ^7,8^. The international monetary fund estimated at the beginning of the pandemic that global growth is expected to fall to -3 percent, making these lockdowns the worst recession since the Great Depression, while the cumulative loss to global gross domestic product (GDP) over 2020 and 2021 was estimated to be around 9 trillion dollars ^9^. Therefore, in face of additional waves of this pandemic, it is important to compare the cost-effectiveness of various strategies to reduce COVID-19 spread, the economic consequences of which might affect the lives of the majority of the world’s population. Here we performed a cost-effectiveness analysis comparing between national lockdowns and the alternative “testing, tracing, and isolation” approach. The latter approach selectively isolates individuals at high exposure risk, thereby minimizing the economic impact. Our findings should be considered by decision-makers during the current and future pandemics.

## Methods

### Model construction

In this study, we applied a modified Susceptible, Exposed, Infectious, Recovered and Deceased (SEIRD) model ^10^, comparing two major strategies: 1. Non-selective nationwide lockdown; 2. Focused isolation of individuals at high exposure risk, who will return to the workforce under social distancing measures after a 14 day isolation period. The model’s variables are compatible with currently established parameters in the literature and our calibration and rely on the actual number of deaths in Israel (according to the ministry of health).

Israel has a population of approximately 9 million. A simulation model of SARS-CoV-2 transmission was constructed (Fig S4). The model includes six compartments (modified SEIRD model ^10^):

1. Susceptible (S): Individuals susceptible to SARS-CoV-2 infection.
2. Exposed (E): Individuals infected with SARS-CoV-2, who can transmit the virus to susceptible individuals. These exposed individuals may be tested for SARS-CoV-2 and isolated accordingly. We further divided the exposed state into two compartments: exposed people who do not develop clinical symptoms (EA) and exposed people who will develop clinical symptoms (I).
3. Exposed asymptomatic (EA): Patients infected with SARS-CoV-2 who do not develop clinical symptoms and can transmit the virus to susceptible patients. These patients may be tested for SARS-CoV-2 and isolated accordingly.
4. Infected (I): Symptomatic SARS-CoV-2 patients that will be completely isolated to prevent further transmission.
5. Recovered (R): Patients who were infected with SARS-CoV-2 and recovered (recovery does not confer lifelong immunity).
6. Death (D): Patients who died because of coronavirus disease 2019 (COVID-19) complications.

The mathematical ordinary differential equations (ODE) that illustrate our model are displayed in Fig S4.

β – transmission rate from carriers (E) to the susceptible population (S). We assume that the transmission rates from the infectious population (I) and from the carrier population (EA) are identical. However, if infectious subjects are isolated, the rate of transmission from the “I” to the “S” population will decrease. If the size of the “I” population decreases by 50% due to isolation, the rate of transmission from “I” to “S” will decrease by 50% (β1β=0×50%). Similarly, if we isolate the “E” population, the transmission rate from “E” to “S” will decrease accordingly (β2).

θ – the proportion of exposed individuals that will develop symptoms.

σ – transition rate from carrier state (E) to infected state (the time from exposure to symptom onset), assuming that the incubation period has an exponential distribution.

δ – mortality rate of the infected population.

γ – recovery rate of the infected population.

α – transmission rate from the recovered population to the susceptible population.

R_0_ (basic reproductive number) – This reflects the number of secondary infections where the whole population is susceptible. If R_0_<1, then the pathogen will be cleared from the population.

Otherwise, the pathogen will be able to infect the whole susceptible population. Note that 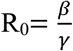. We estimated the number of deaths and the total cost of each strategy and calculated the cost per avoided death (denoted by incremental cost-effectiveness ratio, ICER). Isolation costs for non-infected individuals were based on GDP per capita per day in Israel ($40,270/300 days ^11^), namely $130. Since ~30% of the individuals were working from home during the lockdown in Israel (Eurofund data, ^12^) and 20% of the isolated individuals are retired, we estimated that the cost of isolating one individual per day is $70 (range 50-120, Table 1). The average cost of isolating infected individuals was estimated to be $250 per patient per day (150-350, Table 1). This number sums up: loss of work days $70 (as described above) which applies to all infected individuals. COVID-19 infection is either asymptomatic or causes mild symptoms in most patients. Severe disease typically develops only in ~ 20% of infected individuals, ~5% of which will require intensive care ^13^. Thus, we estimated that ~ 80% of the infected individuals will be isolated at home or in a dedicated facility assuming an average cost of $60 per day (0.8 × $75 isolation costs per day). Hospitalization in Medicine Departments will be required for about 15% of the infected individuals reaching an average cost of $75 per day (0.15 × $500 hospitalization costs per day). While, intensive care unit (ICU) hospitalization will be required for about 5% of the infected individuals and will cost on average $ 45 (0.05 x $900 hospitalization costs). Prices were determined according to the ministry of health publications. The time horizon in the model was 200 days.

**Table 1.**
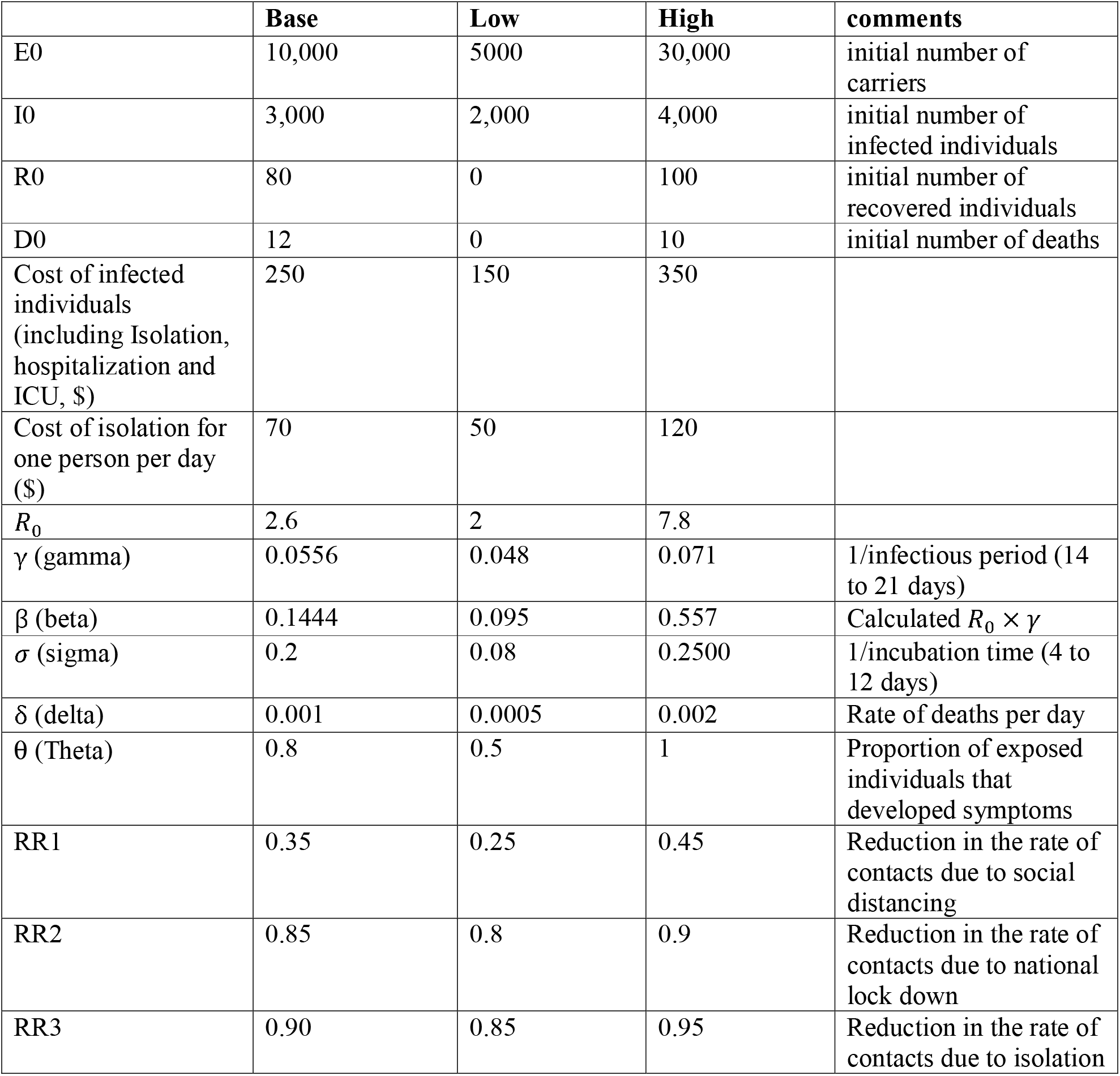
General assumptions including the range for sensitivity analysis.

|  | **Base** | **Low** | **High** | **comments** |
| --- | --- | --- | --- | --- |
| E0 | 10,000 | 5000 | 30,000 | initial number of carriers |
| I0 | 3,000 | 2,000 | 4,000 | initial number of infected individuals |
| R0 | 80 | 0 | 100 | initial number of recovered individuals |
| D0 | 12 | 0 | 10 | initial number of deaths |
| Cost of infected individuals (including Isolation, hospitalization and ICU, \$) | 250 | 150 | 350 | |
| Cost of isolation for one person per day (\$) | 70 | 50 | 120 | |
| $R_0$ | 2.6 | 2 | 7.8 | |
| $\gamma$ (gamma) | 0.0556 | 0.048 | 0.071 | 1/infectious period (14 to 21 days) |
| $\beta$ (beta) | 0.1444 | 0.095 | 0.557 | Calculated $R_0 \times \gamma$ |
| $\sigma$ (sigma) | 0.2 | 0.08 | 0.2500 | 1/incubation time (4 to 12 days) |
| $\delta$ (delta) | 0.001 | 0.0005 | 0.002 | Rate of deaths per day |
| $\theta$ (Theta) | 0.8 | 0.5 | 1 | Proportion of exposed individuals that developed symptoms |
| RR1 | 0.35 | 0.25 | 0.45 | Reduction in the rate of contacts due to social distancing |
| RR2 | 0.85 | 0.8 | 0.9 | Reduction in the rate of contacts due to national lock down |
| RR3 | 0.90 | 0.85 | 0.95 | Reduction in the rate of contacts due to isolation |

### Strategies

We define three levels of preventive measures:

**Social distancing** – maintaining a 2-meter distance between individuals not living in the same household, wearing face masks in public and avoiding gatherings.

**National lockdown (quarantine)** – separating and limiting movement of individuals by confining them to their homes to prevent exposure and infection. Individuals are allowed to leave their home for grocery shopping or medical treatment only. There is no limit on interactions between individuals living in the same household.

**Isolation** – complete isolation of infected individuals or individuals at high exposure risk in a dedicated facility. Home isolation could serve as an alternative by preventing interactions with family members and avoiding the use of shared household items.

We focused on two main strategies (Table S1):

<u>Strategy 1:</u> National lockdown of the susceptible population. Individuals who have essential occupations, as determined by government decisions, will not be quarantined and will be required to maintain social distancing. All known exposed individuals will be completely isolated for a 14-day period according to the standard recommendation by the centers for disease control and prevention (CDC, ^14^) which are also adopted in Israel.

<u>Strategy 2:</u> The susceptible population will be required to maintain social distancing. All known exposed individuals will be isolated. Individuals who are at high exposure risk due to contact with an infected individual or a carrier will be located using detailed epidemiological tracing and/or mobile phone and satellite technology. These people will be isolated for a 14-day period. Under this strategy, only the high exposure risk group will be quarantined, while most of the susceptible population will go back to the workforce under social distancing. In addition to the two main strategies mentioned above, we estimated the number of deaths and the size of the infectious population, without any interventions. The list of parameters in the model as well as the values used for sensitivity analysis are presented in Table 1.

### Parameter estimation and calibration

To calibrate the model, we fitted the observed number of COVID-19 reported deaths between March 27 to June 30, 2020 in Israel, to the number of deaths predicted by the model. We used a simulation annealing method to find the model parameters values. The measure of fitting used the sum of squared errors of the number of deaths obtained by the model output and the observed number of COVID-19 reported deaths during March 27 to June 30, 2020 in Israel. The following parameter estimates were used to construct the model (Table 1):

Exposed (Carriers E0) were defined as the number of infected individuals who are currently undocumented. We determined the estimate of undocumented carriers to 10,000 according to the calibration. This is compatible with a screening conducted in Iceland ^15^, where 0.6-0.8% of the population were carriers and 46-59% have developed symptoms. To validate the model, we compared the observed number of deaths in Israel during April 22 to June 30, 2020 to the number of deaths predicted by the model and observed a nearly perfect fit (Fig S5). The initial number of infected individuals (I0) was determined according to the Israeli Ministry of Health publications on March 27, 2020. The number of susceptible individuals was defined as a rounded approximation of the population in Israel (9 million) after subtracting the infected and undocumented carriers.

An estimation of 18 days was used to calculate the recovery rate (γ) ^16^. The average incubation period (σ) was estimated to be ~ 5.1 days ^13,17,18^.

We used R_0_=2.6 (range 2.0-7.2) ^19^, assuming that the infectious period is 18 days ^16^. Thus the transmission rate from infected (carrier) patients to the susceptible population (β) was 0.144 (range 0.095 to 0.557) ^19^. As all individuals entering Israel from abroad are isolated, we did not include imported cases in the model. Case fatality rates vary between different countries, ranging from 7.2% in Italy to 0.2% in Germany ^20^. Thus, for the rate of death (δ) in our basic model we considered a moderate estimate of 1.8 % or 0.001 per day over 18 days of infection.

RR1-3 represents the reduction rate in contacts due to the implicated preventive measures. The estimation of these parameters was based on the calibration analysis (Fig. S5).

The assumptions used to describe the proportion of the population in compliance with the different levels of preventive measures for each strategy are described in Table S2. For example, we assumed that in strategy 1, the proportion of the susceptible population under lockdown (r_S2) will be 80%, and the proportion of the carrier population that is under isolation in both strategies (r1_E, r2_E) was estimated at 30%. The time horizon was 200 days and therefore the discount rate was 0. In addition, we assumed that the cost of isolation in strategy 1 is only for 28 days.

The analysis was performed using MATLAB.

### Sensitivity analysis

One-way sensitivity analysis was used for all the parameters in the model (see Table 1 and Table S2 for the range used for the sensitivity analysis). Tornado diagram was constructed for the ICER and the number of deaths in each strategy.

### Probabilistic sensitivity analysis

A probabilistic sensitivity analysis (Monte Carlo simulation) was performed to assess the effects of ranging base case variables on the model outcomes with 1000 draws from probability model parameters.

### Ethical considerations

The study has received an IRB exempt from the local institute (Rabin Medical Center).

## Results

We first tested our model with strategy 1: National lockdown, isolation of all infected individuals and a 14-day isolation period for the high exposure risk group. Applying this strategy will yield a peak of 7,671 infected individuals followed by a rapid decline (Fig. 1, right upper panel) with a total of 9,046 infected individuals over the 200-day period. Under this strategy, the number of expected deaths is 303.5 (Fig. 1, right lower panel). This number will be reached after ~110 days. In strategy 2, infected individuals will remain under isolation. However, in strategy 2, instead of a national lockdown, only high exposure risk individuals will be isolated. Following 14 days of isolation, these individuals will return to the workforce while maintaining social distancing. Under these conditions, the maximum number of infected individuals will slightly increase to 19,646, with a total of 38,761infected individuals over the 200 days period (Fig. 2, right upper panel). The total number of deaths will increase to 577.8 (Fig. 2, right lower panel).

**Figure 1:**
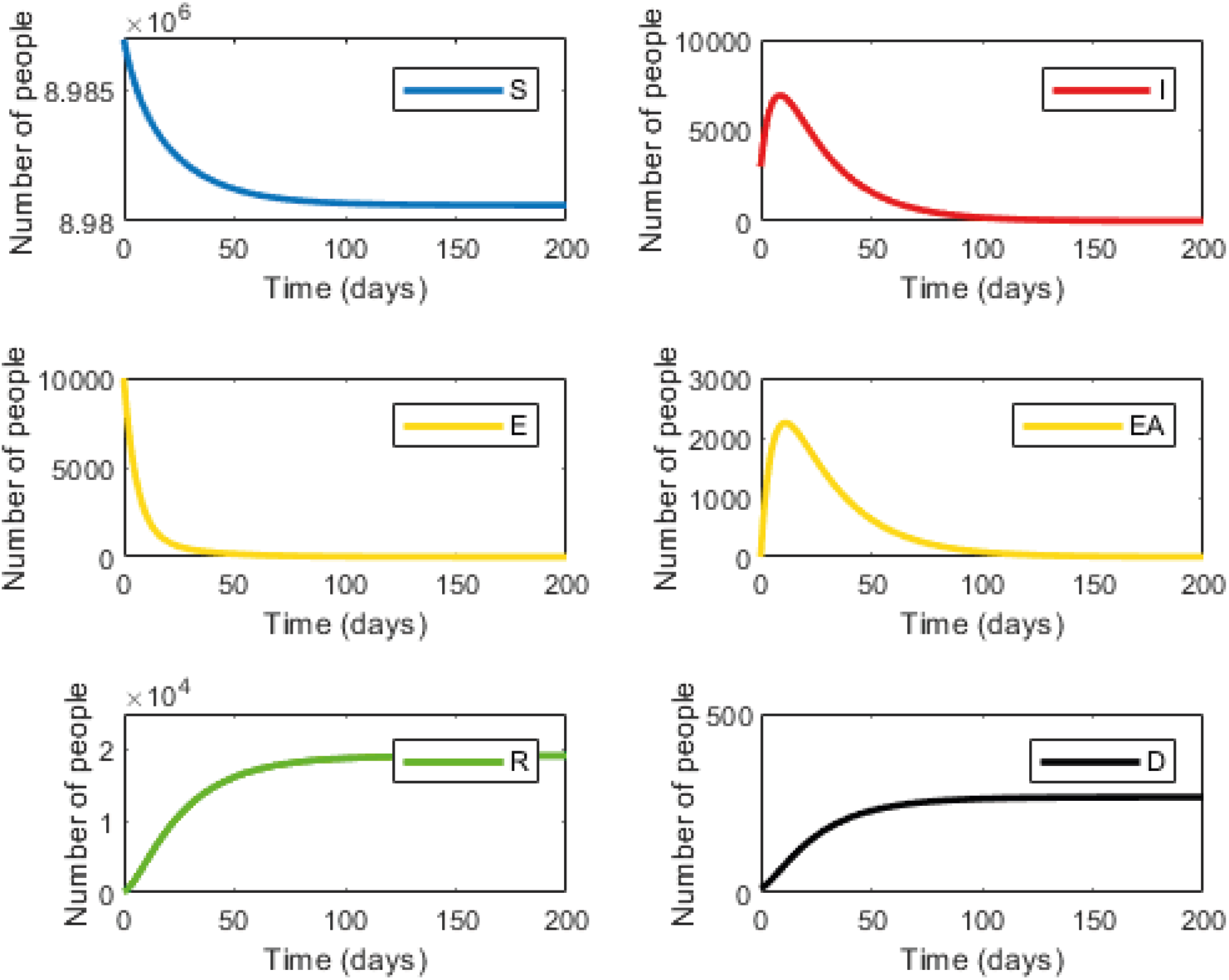
Infection dynamics under national lockdown (strategy 1) The graphs display the dynamics within the six compartments over time. Susceptible (S, blue); Infected (I, red); Carrier (Yellow, E); Carrier asymptomatic (Yellow, EA); Recovered (R, Green), Dead (D, Black).

**Figure 2:**
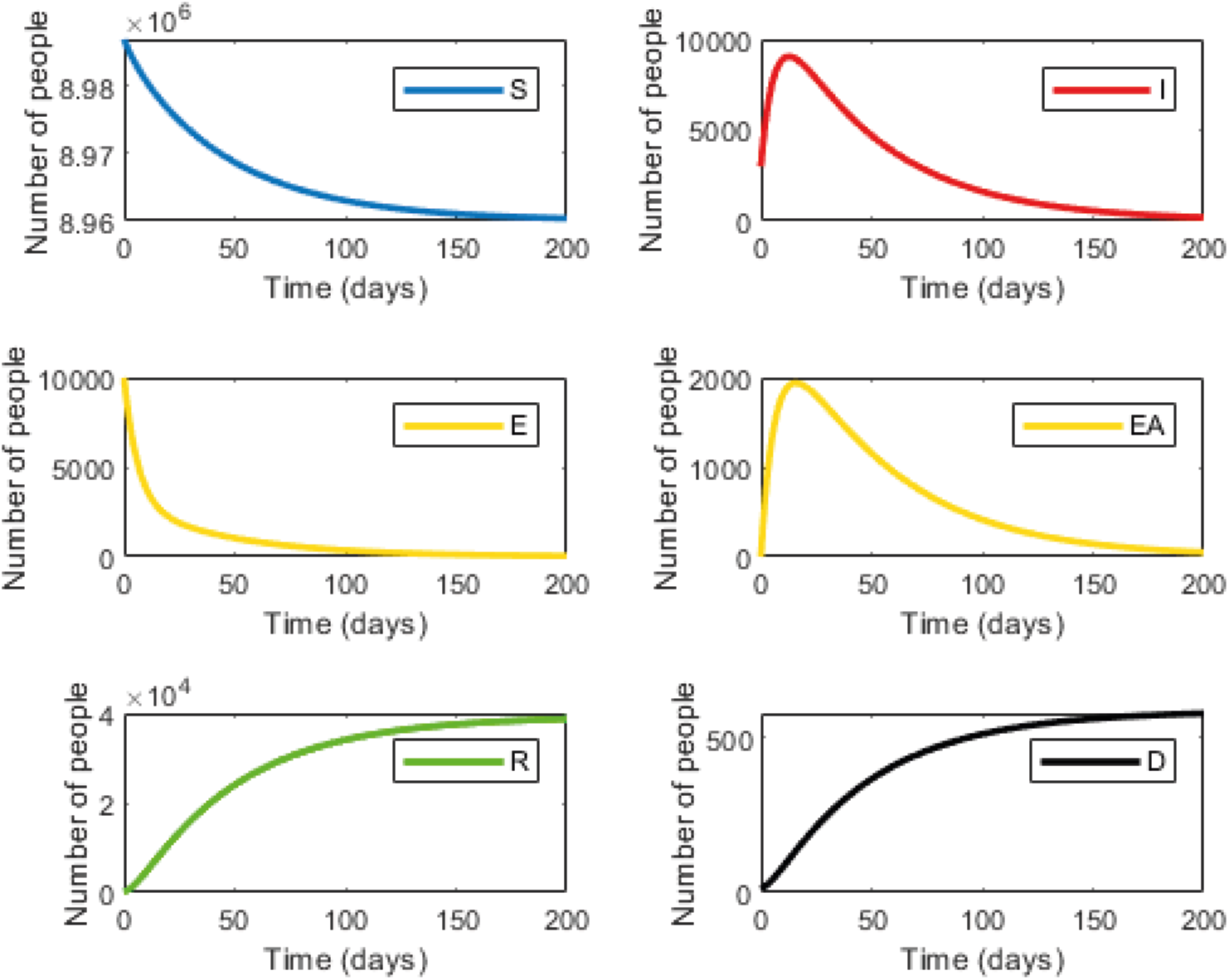
Infection dynamics during isolation of individuals at high-risk of exposure (strategy 2) Graphs are labeled as described in Figure 1.

Table S3 summarizes the costs and the number of expected deaths under each strategy. Factoring the cost of lost workdays and isolation enabled us to calculate the incremental cost. The incremental cost divided by the incremental number of deaths yields the incremental cost-effectiveness ratio (ICER). The calculated cost for strategy 1 was $12,495 million and for strategy 2 $122.9 million. While the expected number of deaths was 303.5 for strategy 1 and 577.8 for strategy 2. Thus, incremental cost would be $ 12,372 million and the incremental number of deaths would be 274, while the resulting ICER would be $45.1 million. Thus, under these conditions, the cost of preventing one death case would be $ 45,104,156.

One quality-adjusted life year (QALY) equals one year of healthy life, and since the majority of patients who succumbed to the COVID-19 were older non-healthy individuals (mean age ~80 in Israel with a life expectancy of 8.56 years), we estimated that on average, every death case will result in a loss of 10 QALYs. Assuming that one case of death is equivalent to a loss of 10 QALYs, the ICER would be $45,104,156 divided by 10, resulting in $4.5 million per QALY. This number is much higher than the internationally accepted ICER threshold values (between $50,000-150,000 per QALY^21^). Furthermore, this number is higher than the $100,000-150,000 per QALY, recommended by Neumann et al. as the threshold of willingness to pay ^22^. Notably, in Israel, the ICER threshold value is estimated at around $15,243 – 17,366 per QALY^23^.

Another parameter used to assess the cost of death in other fields, is the value of a statistical life (VSL), a statistical measure of the willingness to pay for small reductions in mortality risks, estimated to be ~$ 10,000,000 ^24^. Similarly, this value is much lower than our calculated ICER. The results of a one-way sensitivity analysis for both strategies are displayed in Figure 3. The virus transmission rate (β) and the daily mortality rate (δ) were among the variables with the strongest influence on ICER (Fig 3). A one-way sensitivity analysis showing the effect of these parameters on ICER is displayed in Fig S1.

**Figure 3:**
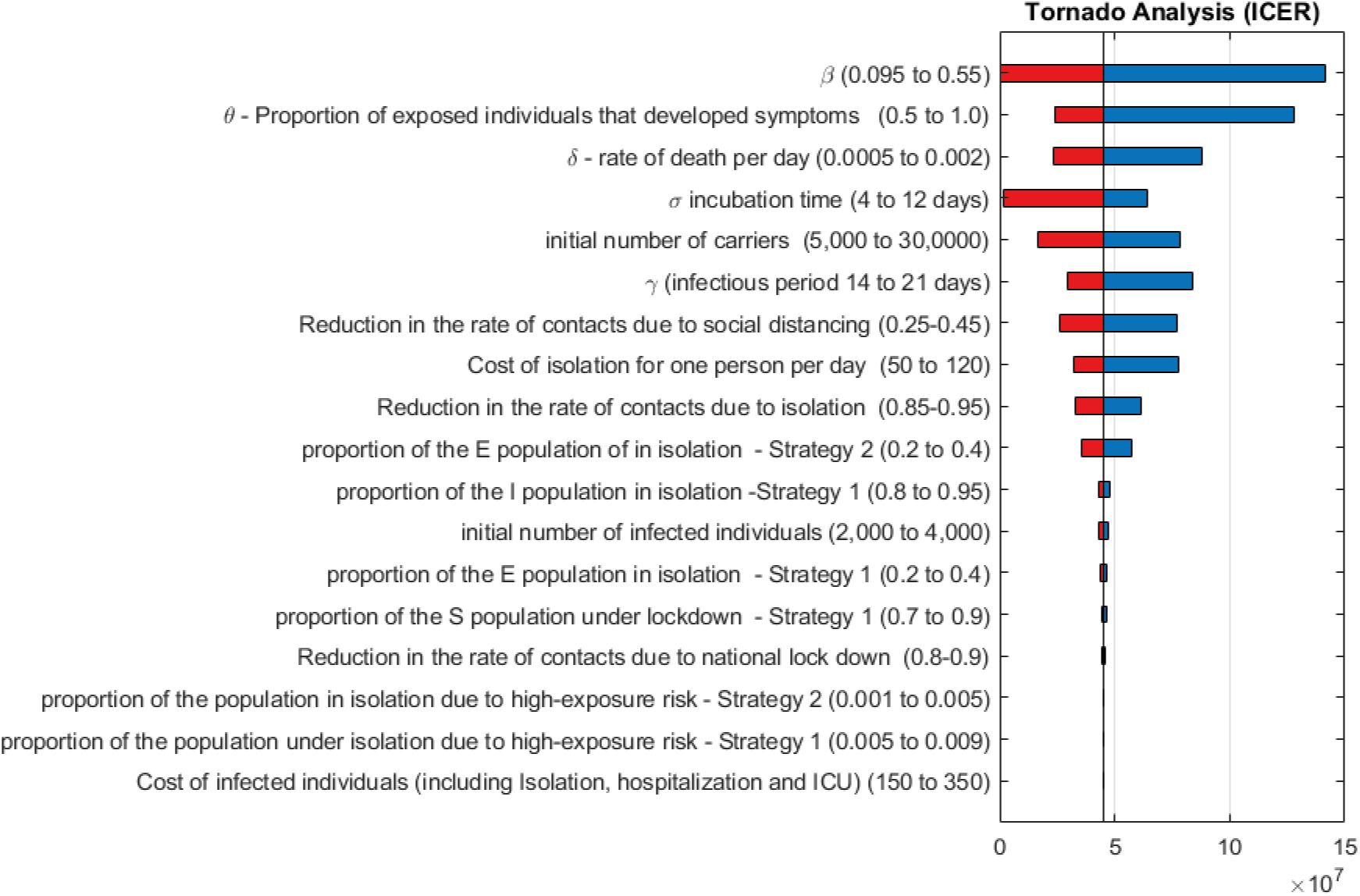
Sensitivity analysis (Tornado diagram) of the incremental cost-effectiveness ratio (ICER) between the two strategies. Red bars indicate that the value was produced by the Lower Bound (Low), and dark blue bars indicate that the value was produced by the Upper Bound (High).

An additional sensitivity analysis was performed for the number of expected deaths under both strategies. As shown in Figure S2, the initial carrier population, the daily mortality rate (δ) and the virus transmission rate (β) were the parameters with the greatest effect on the death rate. A probabilistic sensitivity analysis of the cumulative death for each strategy and of the estimated distribution of the ICER are displayed in Figure 4 and Figure 5, respectively. The median difference between strategy 1 and 2 in the number of deaths is 124 (IQR: 71-221) and the median ICER is $ 49.6 million (IQR: 22.7-220.1 million).

**Figure 4:**
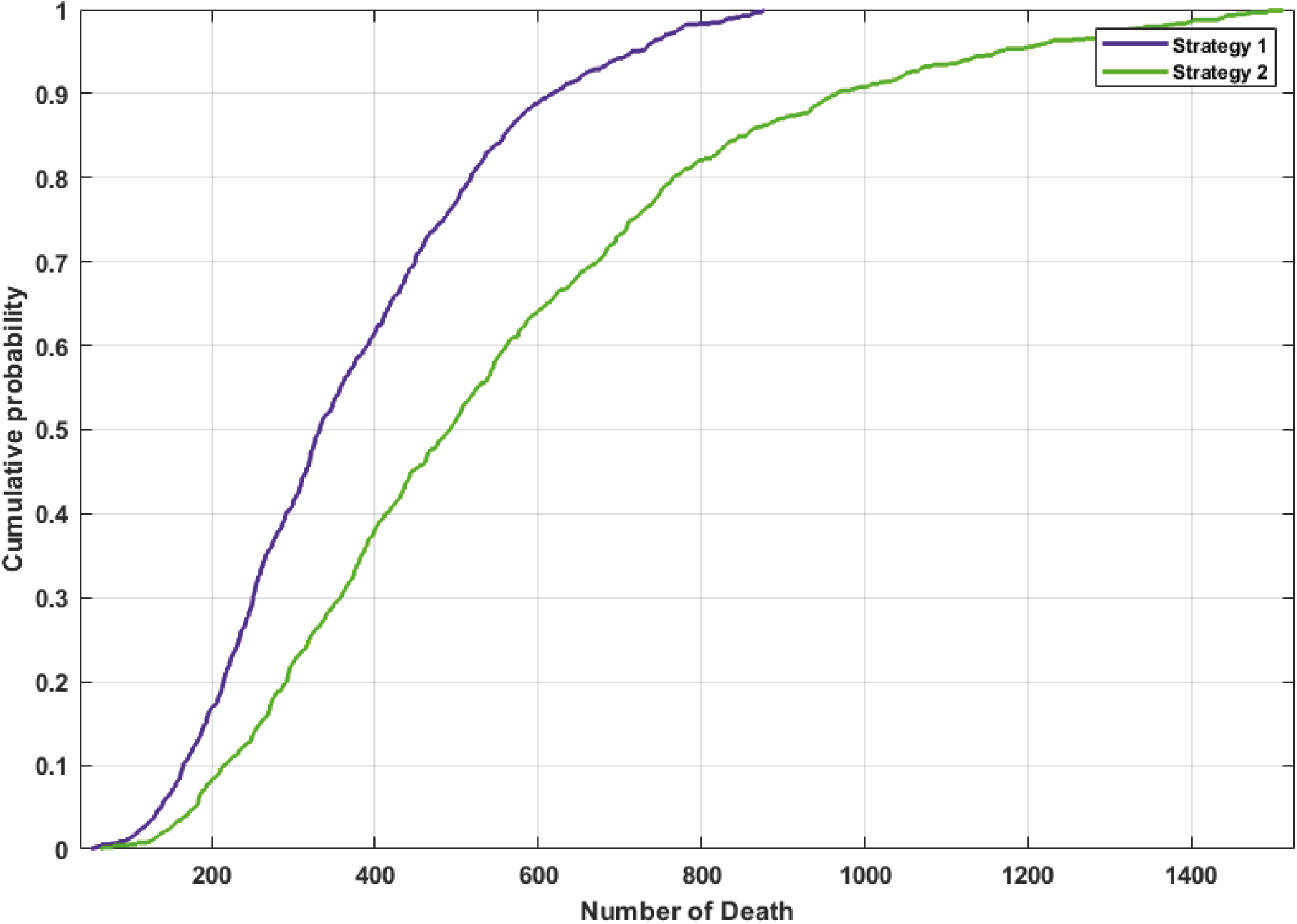
Probabilistic sensitivity analysis for the number of deaths in each strategy. A probabilistic sensitivity analysis was used to assess the probability distribution of the expected number of deaths according to each strategy.

**Figure 5:**
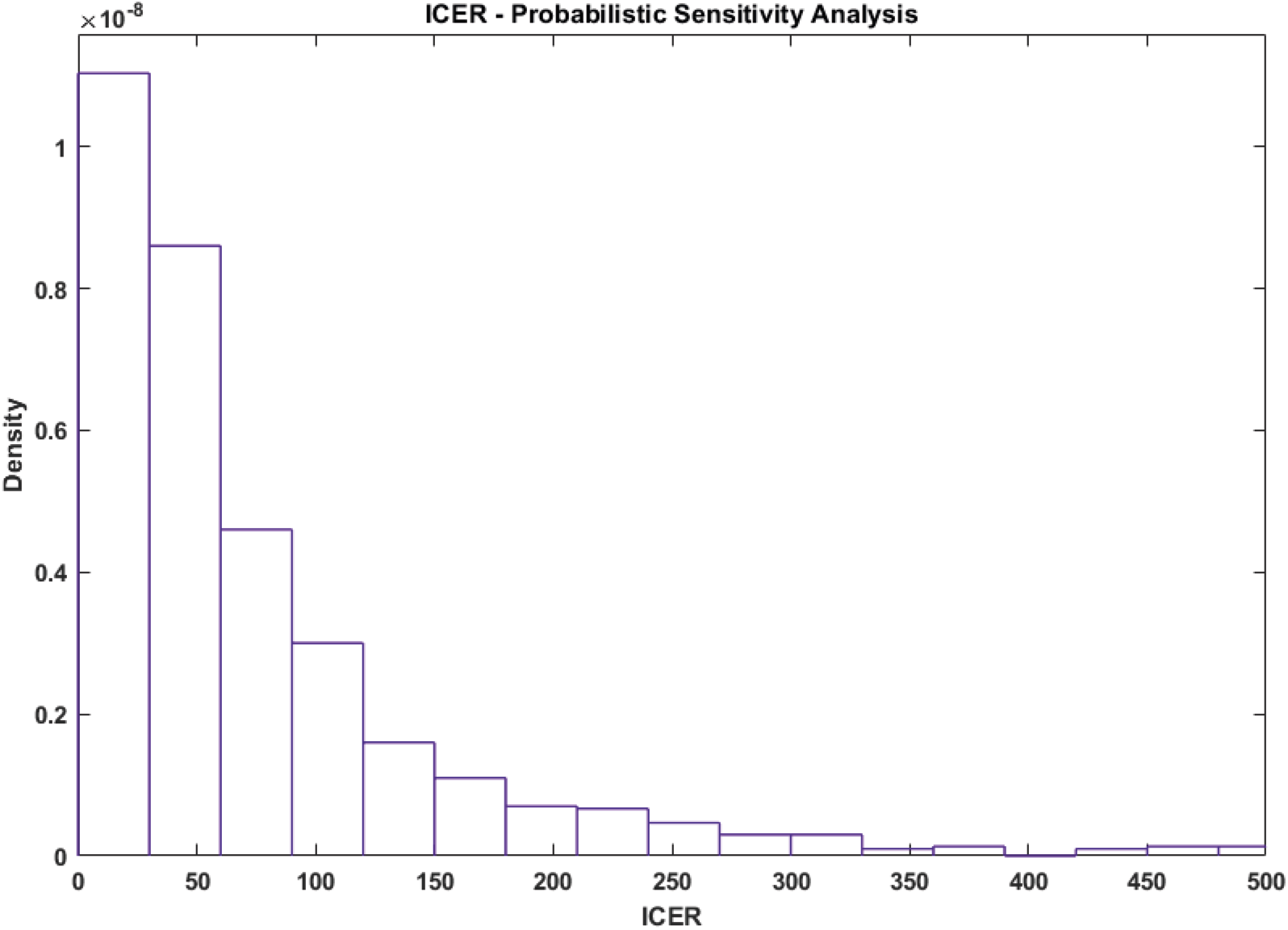
Probabilistic sensitivity analysis for the ICER. A probabilistic sensitivity analysis was used to assess the probability distribution of the ICER (expressed in millions of US $). The graph was truncated at $500 million.

Finally, we tested our model without any intervention. Starting from a baseline of 10,000 carriers and 3,000 infected individuals, the maximal number of infected individuals is predicted to be 1.94 million (Fig. S3, right upper panel). This number will be reached after 200 days. Under these conditions, the expected number of deaths will be 118,130 (Fig. S3, right lower panel).

## Discussion

In this study, we applied cost-effectiveness analysis tools to distinguish between two different strategies aimed to slow down the virus spread. We show that a national lockdown (strategy 1) will result in a total of ~ 19,646 infected individuals and around 303 deaths over a period of 200 days. An alternative “testing, tracing, and isolation” approach (strategy 2) in which only individuals with a high exposure risk are isolated will result in a total of 38,761 infected individuals and 577.8 deaths. Overall, strategy 1 is expected to save ~274 more lives, but with a cost of $ 45.1 million to prevent one death case, compared to the more focused approach.

One of the major concerns during a pandemic is that the number of patients needing intensive care and mechanical ventilation will overwhelm the local health care system. According to our model, a no-intervention approach will result in an unaccepTable number of infected individuals (1.94 million) and an extremely high death rate (~ 118,130), and therefore is not realistic. In contrast, the difference in the peak number of infected patients between the two main strategies tested is ~1,375 patients, in favor of strategy 1. Assuming that ~20% of the patients are at risk of developing a severe form of the disease ^13^, this translates into an additional 275 potential hospitalizations and mechanical ventilations at the peak, which is an acceptable burden.

As with similar models, the validity of this model is based on the correct assumption of the various parameters. Since SARS-CoV-2 is a new virus and the exact infection rates in the population are still unknown, these parameters are subject to variations and may change over time. To validate our model, we calibrated it by comparing the rate of deaths according to our model assumptions with the real-time published numbers of these parameters in Israel (Fig. S5). The obtained graphs were strikingly similar, further confirming our model.

One major limitation of compartment models is that they assume a homogeneous population, which is not the case in many countries. In Israel, for example, the reproductive number (R_0_) is most probably higher among some religious groups. This might alter the infection dynamics and affect the outcome of the model. If R_0_ is indeed significantly higher in these specific populations, the average R_0_ used in the model will increase. This implies higher infection and death rates. However, if this is indeed the case, specific lockdown measures could be applied to these “red zones”. However, since exact data regarding the R_0_ in these populations is not available, our model does not consider areas with different R_0_ within the country.

Our sensitivity analysis indicates that the transmission rate (β) is the parameter with the most influence on ICER. β relies on R_0_, which might change, and on the infectious period, which will most likely remain the same. However, since a similar R_0_ is used for modeling both strategies, we believe that the difference in the number of deaths and ICER will remain similar.

Obviously, strategy 2 of “trace, isolate, test and treat”, largely represents the steps successfully undertaken by South Korea to handle the situation, demands two critical components:

1. Intensive epidemiological investigations of infected patients combined with surveillance to trace possible interactions. 2. Availability of enough testing kits and facilities to enable a large number of daily PCR tests to isolate subjects at high exposure risk.

Obviously, the intensive epidemiological investigations, necessary to define people at high exposure risk who must be isolated, implies massive use of mobile phone and satellite technology, thereby violating citizens’ privacy rights. Therefore, decision-makers must carefully weigh their options, keeping in mind the very high economic price associated with national lockdown strategies.

In summary, in this cost-effectiveness analysis, we show that over time a strategy of national lockdown is moderately superior to a strategy of focused isolation in terms of reducing death rates, but involves extremely high economic costs to prevent one case of death. These economic costs might add to the future economic consequences of this pandemic, and thus these options should be carefully considered and balanced.

## Data Availability

Data available within the article or its supplementary materials

